# SARS-CoV-2 Detection Using an Isothermal Amplification Reaction and a Rapid, Inexpensive Protocol for Sample Inactivation and Purification

**DOI:** 10.1101/2020.04.23.20076877

**Authors:** Brian A Rabe, Constance Cepko

## Abstract

As the current SARS-CoV-2 pandemic spreads, the need for more diagnostic capabilities is great. In order to address this need, we have developed a highly sensitive RT-LAMP assay compatible with current reagents, that utilizes a colorimetric readout in as little as 30 minutes. In addition to this, we have developed an inexpensive pipeline to further increase sensitivity without requiring highly specialized equipment. A rapid inactivation protocol capable of inactivating virions, as well as endogenous nucleases, was also developed to increase sensitivity and sample stability. This protocol, combined with our RT-LAMP assay, has a sensitivity of at least 50 viral RNA copies per microliter in a sample. To further increase the sensitivity, a purification protocol compatible with this inactivation method was developed. The inactivation and purification protocol, combined with our RT-LAMP assay, brings the sensitivity to at least 1 viral RNA copy per microliter in a sample. We hope that this inactivation and purification pipeline, which costs approximately $0.07 per sample and which uses readily available reagents, will increase the availability of SARS-CoV-2 testing, as well as expand the settings in which this testing can be performed.

## Introduction/Background

The current SARS-CoV-2 pandemic has had and will continue to have an enormous impact on society worldwide, threatening the lives and livelihoods of many. As the disease spreads, the need for rapid point-of-care diagnostic tools has become immense. Many efforts are currently underway to develop such an assay that can be easily used in a variety of settings^1^. The ideal assay would require no specialized equipment and would have a rapid and easily read result. To that end, we have developed an assay based upon the reverse-transcription loop-mediated isothermal amplification (RT-LAMP) technique. To boost sensitivity, we also developed a novel sample preparation method that can be used as the first step for many different types of downstream assays. The protocol is simple, inexpensive, and rapid. It utilizes reagents that are readily prepared in large quantities.

LAMP is a method of isothermal DNA replication that utilizes, in an accelerated format, six DNA oligos that hybridize with eight different regions of a target molecule^2^. Utilizing a strand displacing polymerase and loops formed during this reaction, a fast amplification reaction can occur upon proper oligo binding to the desired target. Such reactions are capable of generating microgram quantities of DNA in a very short period of time at a single reaction temperature. Furthermore, although the strand-displacing polymerase has reverse transcriptase activity, a reverse transcriptase can be included to improve sensitivity within the reaction when detecting an RNA target (RT-LAMP), such as the SARS-CoV-2 RNA. LAMP assays have a variety of readouts due to the large amount of DNA generated, including fluorescence using an intercalating DNA dye, turbidity, or by a drop in the pH if the reaction is minimally buffered^1, 3, 4^. This change in pH, sufficient to cause a pH indicator dye to visibly change color, is the most applicable method for a point-of-care LAMP-based diagnostic.

We decided to design and test our own RT-LAMP assay utilizing the LAMP reaction reagents from New England Biolabs (NEB). For each of 11 assays tested we used PrimerExplorer V5 (https://primerexplorer.jp/e/) to design all primers, with the exception of the loop primers for Assay 1. PrimerExplorer could not find a set for this region of the genome, so Assay 1 primers were designed manually. As we prepared to test these assays, we learned of several other assays designed by researchers at NEB, and so we also tested their two most sensitive assays, Gene N-A and Orf1a-C, in order to compare with our own assays^1^.

One of our assays, HMS Assay 1, performed particularly well compared to the others (data not shown for others). We then modified the forward inner primer (FIP) and backward inner primer (BIP) of this assay to include a “TTTT” linker between the F1c and F2 regions to create HMS Assay 1e, as this has been reported to further improve the reaction (for HMS Assay 1 and HMS Assay 1e oligo sequences, see Table 1)^5^. HMS Assay 1/1e is designed within the ORF1ab of SARS-CoV-2 in a region that is not highly conserved with either SARS or Bat SARS-like coronavirus isolate Rs4084, two closely related coronaviruses (Figure 1). As we demonstrate, HMS Assay 1 and HMS Assay 1e outperform NEB Gene N-A and NEB Orf1a-C in both sensitivity and speed.

**Table 1.**
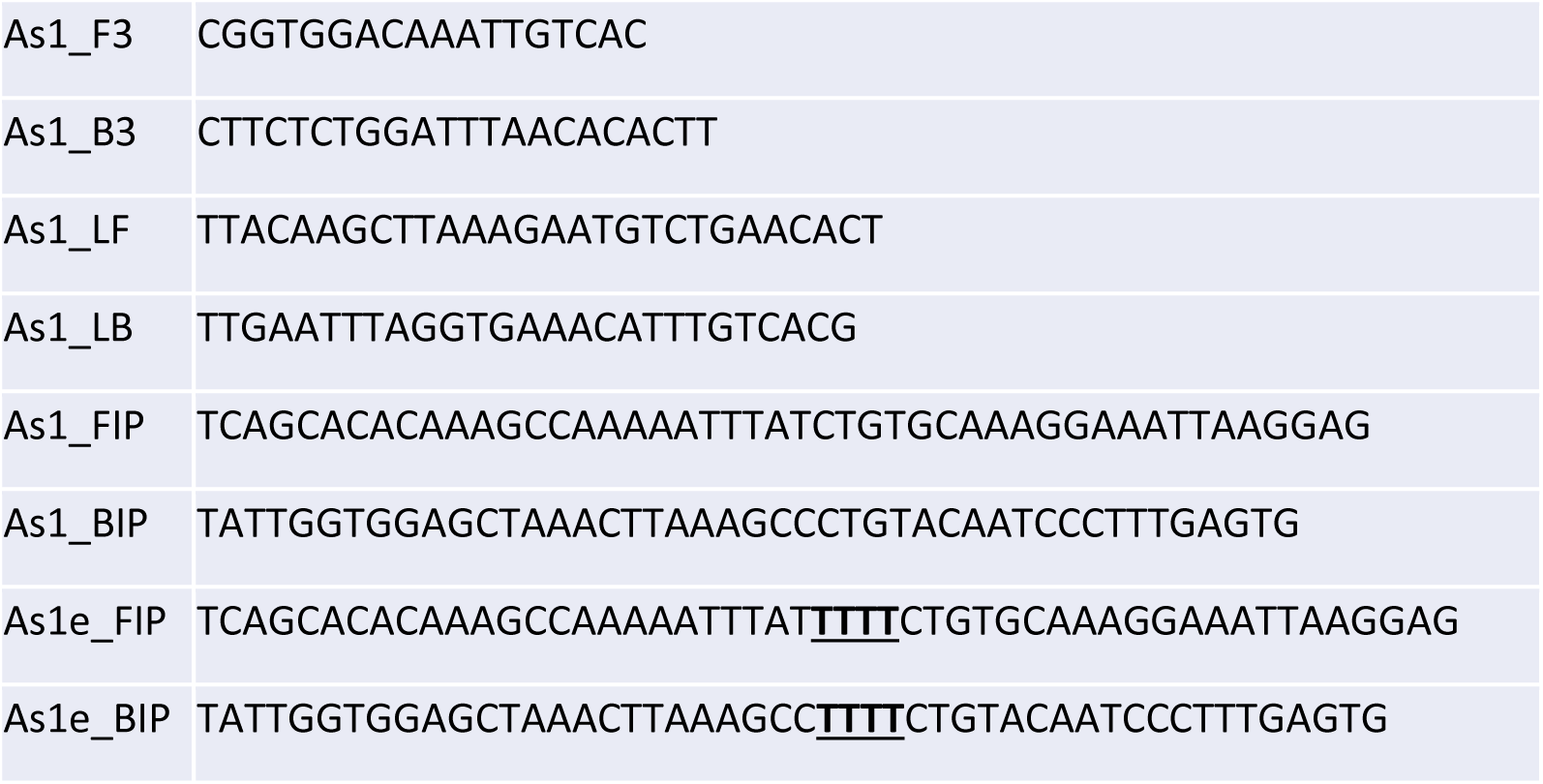
Oligo Sequences used for HMS Assay 1 and HMS Assay 1e. Both HMS Assay 1 and HMS Assay 1e use the same F3, B3, LF, and LB oligos as shown. HMS Assay 1e uses its own FIP and BIP oligos which are identical to those used by HMS Assay 1 with the exception of 4 thymidine residues inserted in the middle (underlined and bold)

**Figure 1.**
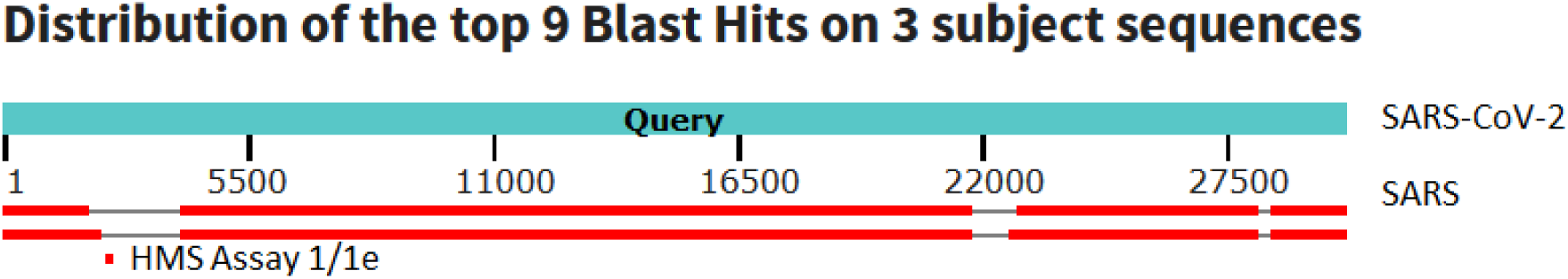
Alignment of SARS-CoV-2 to Related Coronaviruses. An alignment (blastn, megablast) of SARS, Bat SARS-like coronavirus isolate Rs4084, and the sequence detected by HMS Assay 1/1e.

In addition to developing a robust RT-LAMP primer set, we also sought to optimize sample preparation in a way that would maximize sensitivity and render samples stable and safe for those conducting tests. In order to do this, we explored the tolerance of the RT-LAMP reaction to detergents and chaotropic salts that might aid in lysis of virions and purification of viral RNA genomes and mRNA. In addition, we optimized a very simple and rapid protocol for inactivating virions as well as endogenous RNAses in a way that allows at least 5 µl of sample (swabs in saline/PBS or straight saliva) to be added to a reaction and to bring sensitivity to 10-50 RNA copies per microliter of sample. We also developed a simple and rapid process by which viral RNA can be concentrated from as little as 0.5 ml of collection media such that, when used with the HMS Assay 1e, the limit of detection falls at least to 2 RNA copies per microliter. Unlike purification schemes used for the current FDA approved qRT-PCR-based test, this purification does not require a commercial kit or, for most sample types, a centrifuge. It binds nucleic acids to silica in the form of a suspension known as “glass milk”^6^, which is readily available in industrial quantities at little expense. The glass milk purification is completed in one or two tubes, one of which can be the same one in which the reaction is run. The RNA is kept in sodium iodide (NaI) or high ethanol solutions throughout. Both solutions inhibit RNAse activity, ensuring RNA stability. The glass milk preparation for millions of purifications can be made in an afternoon, with no need for specialized equipment or expensive reagents. The overall cost of the inactivation and purification is approximately $0.07 per sample.

## Results

### Sensitivity of HMS and NEB RT-LAMP Assays

We first tested HMS Assay 1, HMS Assay 1e, NEB Gene N-A, and NEB Orf1a-C using the NEB’s WarmStart LAMP Kit (NEB E1700) with a real-time fluorescence-based readout (Figure 2). In this reaction scheme, each cycle represents 30 seconds at 65°C. Ideally, a positive result will be read after 30 minutes, or the 60^th^ cycle, a timepoint used by Zhang et al^1^. We used positive control RNAs from Twist Bioscience (Sku 102019). We ran 10 µl reactions in triplicate, including 0, 100, 200, or 300 viral RNA copies per reaction. As can be seen, all four assays are capable of detecting viral RNA at low copy number, although in this setup NEB Gene N-A demonstrated lower sensitivity and slower amplification than the rest.

**Figure 2.**
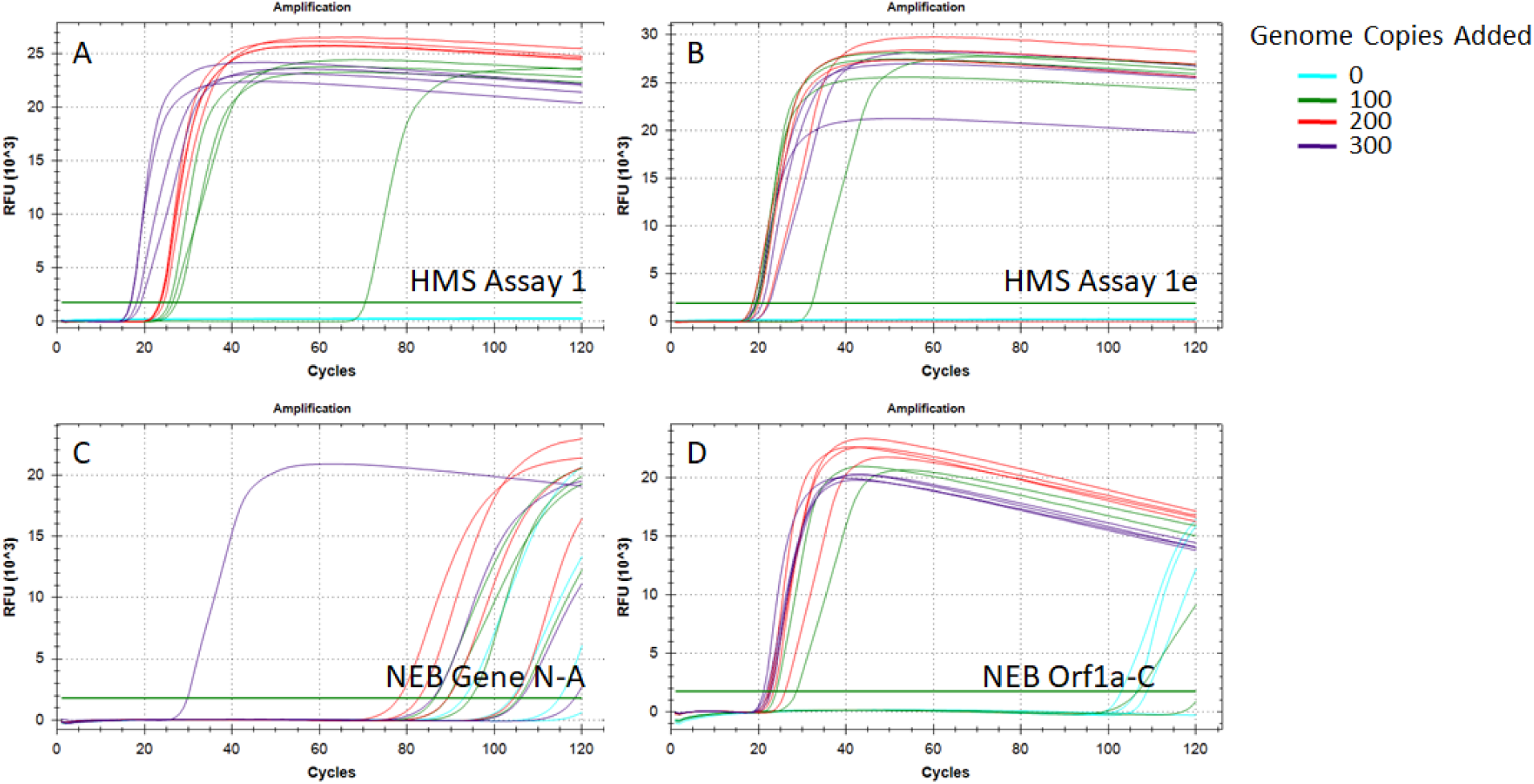
Initial Sensitivity Test of Promising RT-LAMP Assays. 10 µl RT-LAMP reactions were run with a fluorescent readout. 0 (blue), 100 (green), 200 (red), or 300 (purple) control RNAs included per reaction (n = 4). Assays performed were A - HMS Assay 1; B - HMS Assay 1e; C - NEB Gene N-A; D - NEB Orf1a-C. Each cycle corresponded to 30 sec at 65°C, total time run was 60 minutes. RFU – Relative Fluorescence Units

In order to further assess sensitivity, we ran repeated reactions using the same fluorescence-based readout with HMS Assay 1, HMS Assay 1e, and NEB Orf1a-C (Figure 3). For each, we ran 48 10-µl reactions with 200 viral RNA copies each and 48 10 µl reactions with no viral RNA added. As can be seen, both HMS assay 1 and HMS assay 1e performed well, showing high amplification in 45 and 47 out of 48 reactions with 200 RNA copies, respectively. Furthermore, none of the reactions without viral RNA exhibited any amplification by 60 minutes. NEB Orf1a-C did not perform as well, as the time to amplification in the 200 RNA copy reactions was highly variable with many not amplifying until just before or after the 30-minute point. Furthermore, two reactions without viral RNA exhibited amplification, but we cannot rule out the possibility that these reactions, as sensitive as they are, were contaminated. These data suggest that HMS Assay 1 and HMS Assay 1e are the more robust primer sets for this assay.

**Figure 3.**
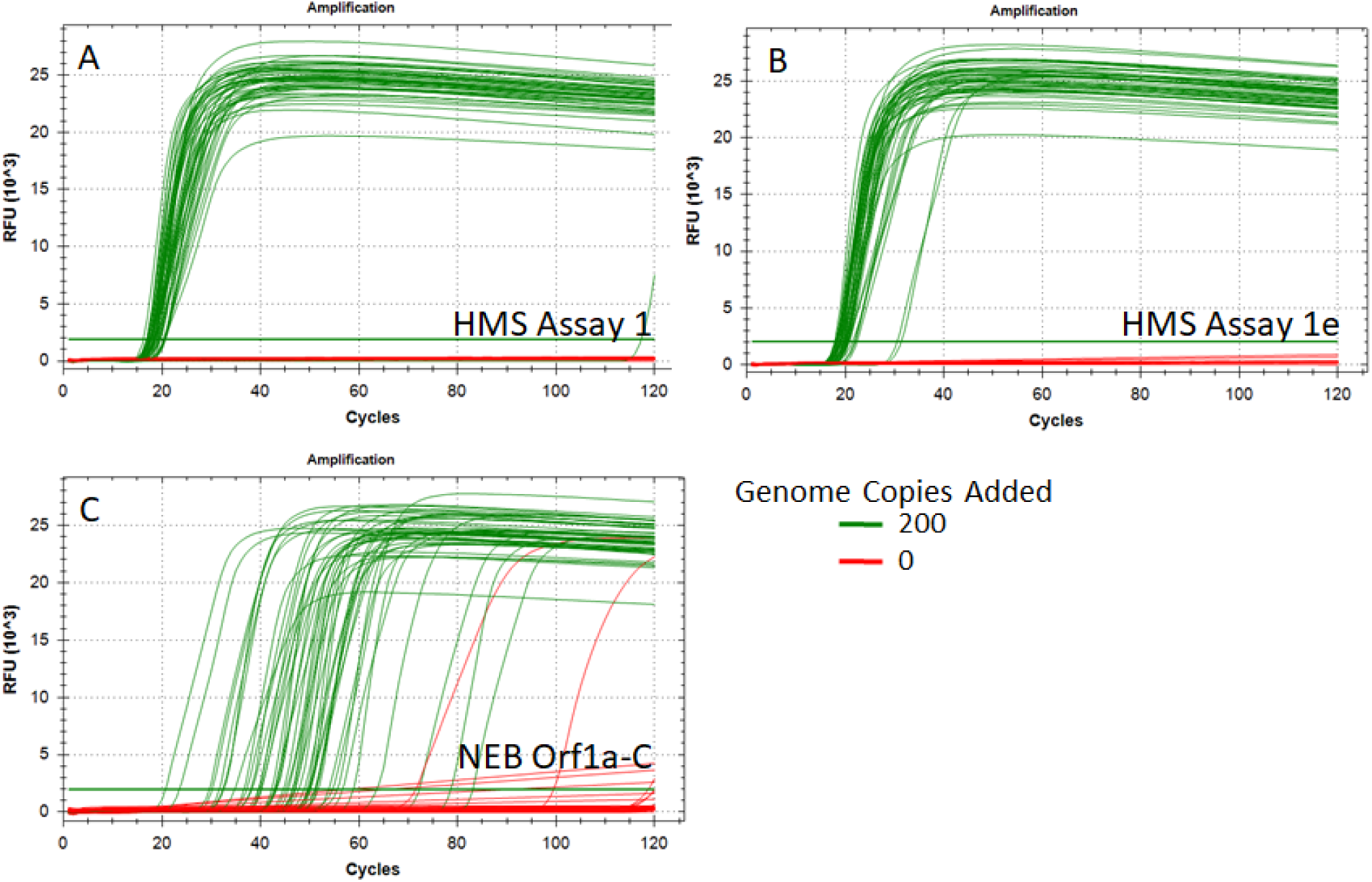
Repetitions of Low Genome Number Reactions. 10 µl RT-LAMP reactions were run with a fluorescent readout. 200 (green) or zero (red) control RNA copies included per reaction (n = 48). Assays performed were A - HMS Assay 1; B - HMS Assay 1e; C - NEB Gene N-A; D - NEB Orf1a-C. Each cycle corresponded to 30 sec at 65°C, total time run was 60 minutes. RFU – Relative Fluorescence Units

### Detergent Tolerance

In order to potentially improve the sensitivity of the RT-LAMP reaction when using patient samples, we hypothesized that an increase in detergent within the reaction might help to lyse virions, making their genomic RNA more accessible for reverse transcription and amplification. Using HMS Assay 1 and the same 10 µl fluorescent reactions as described above, we ran reactions with 500 viral RNA copies and differing amounts of added Tween20 or TritonX100. The reaction is quite tolerant of added detergents, and robust amplification could be seen up to at least 1.5% Tween20 and 1% TritonX100 (Figure 4). Amplification could still be detected for both detergents up to 3%, but the reactions appeared to plateau at a lower level of fluorescence when detergent levels increased.

**Figure 4.**
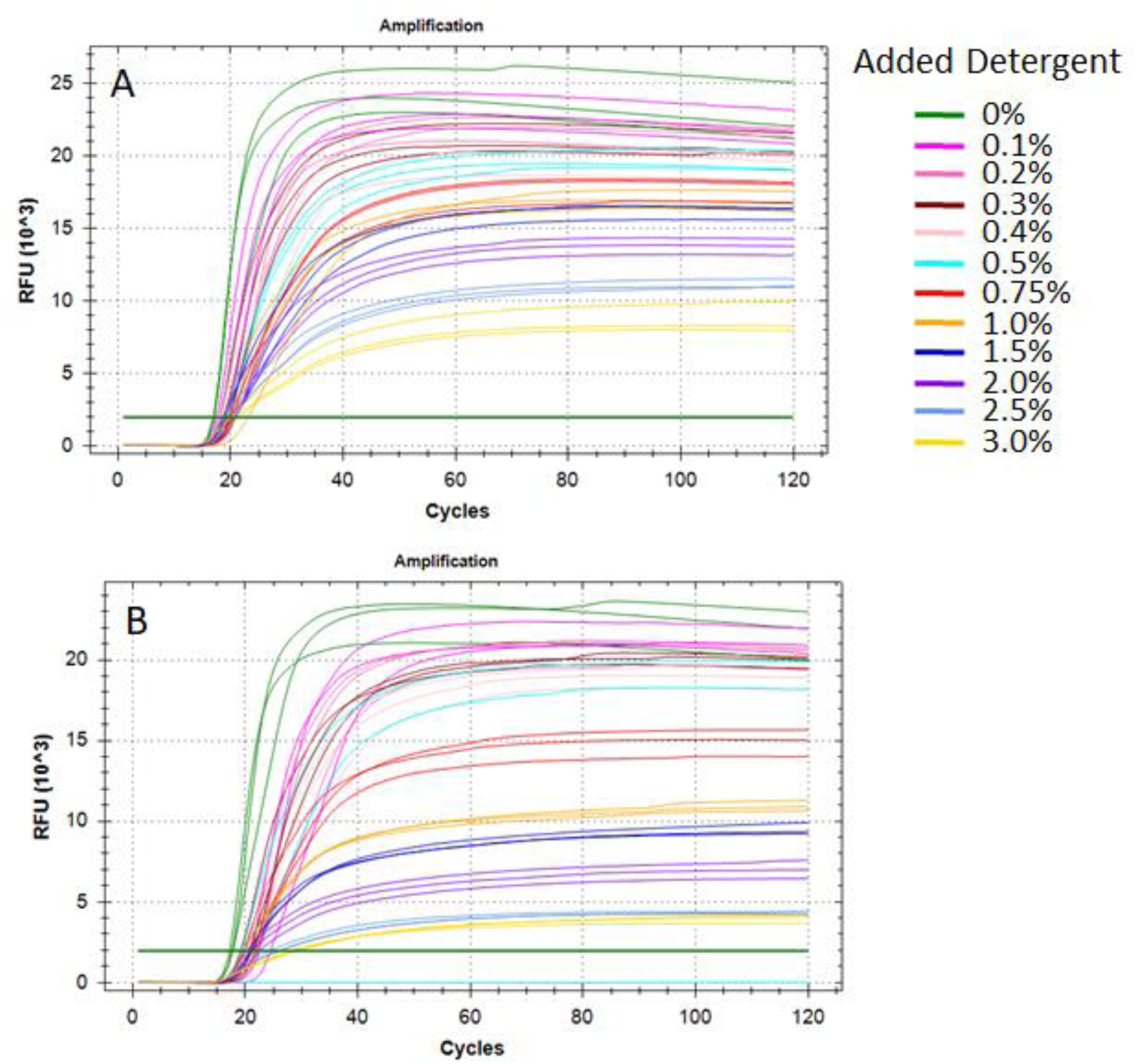
Assessment of RT-LAMP Detergent Tolerance. 10 µl RT-LAMP reactions were run with a fluorescent readout. All reactions contained 500 control RNA copies, the HMS Assay 1 primer set and 0% - 3% added detergent (see legend). Each cycle corresponded to 30 sec at 65°C, total time run was 60 minutes. RFU – Relative Fluorescence Units

### Chaotropic Salt Tolerance

Most RNA purification schemes utilize guanidinium thiocyanate (GuSCN)^7^. This chaotropic salt is a powerful protein denaturant that can aid in lysis and RNAse inactivation. We tested the tolerance of the RT-LAMP reactions to this chemical in order to optimize a rapid purification protocol that might not remove all GuSCN traces. We first created a sample lysis buffer containing 4 M GuSCN and 2% TritonX100. We then added varying amounts of this buffer into HMS Assay 1e 25 µl colorimetric RT-LAMP reactions (NEB M1800) with 500 viral RNA copies, such that the final GuSCN concentration would range from 160 mM to 27 mM (i.e. a final dilution of 1:25 to 1:150 in the final reaction). All colorimetric reactions were run for 30 minutes at 65°C. As Figure 5A shows, the reaction was visibly positive at GuSCN concentrations at or below 53 mM (1:75 dilution) although the 53 mM reaction was slightly more orange than the rest. To repeat this result, we ran eight replicates of the same reactions at 50 mM and 40 mM GuSCN (1:80, and 1:100 dilution, respectively, Figure 5B), and all were robustly positive. This indicates that the colorimetric RT-LAMP reactions are tolerant to GuSCN up to 50 mM.

**Figure 5.**
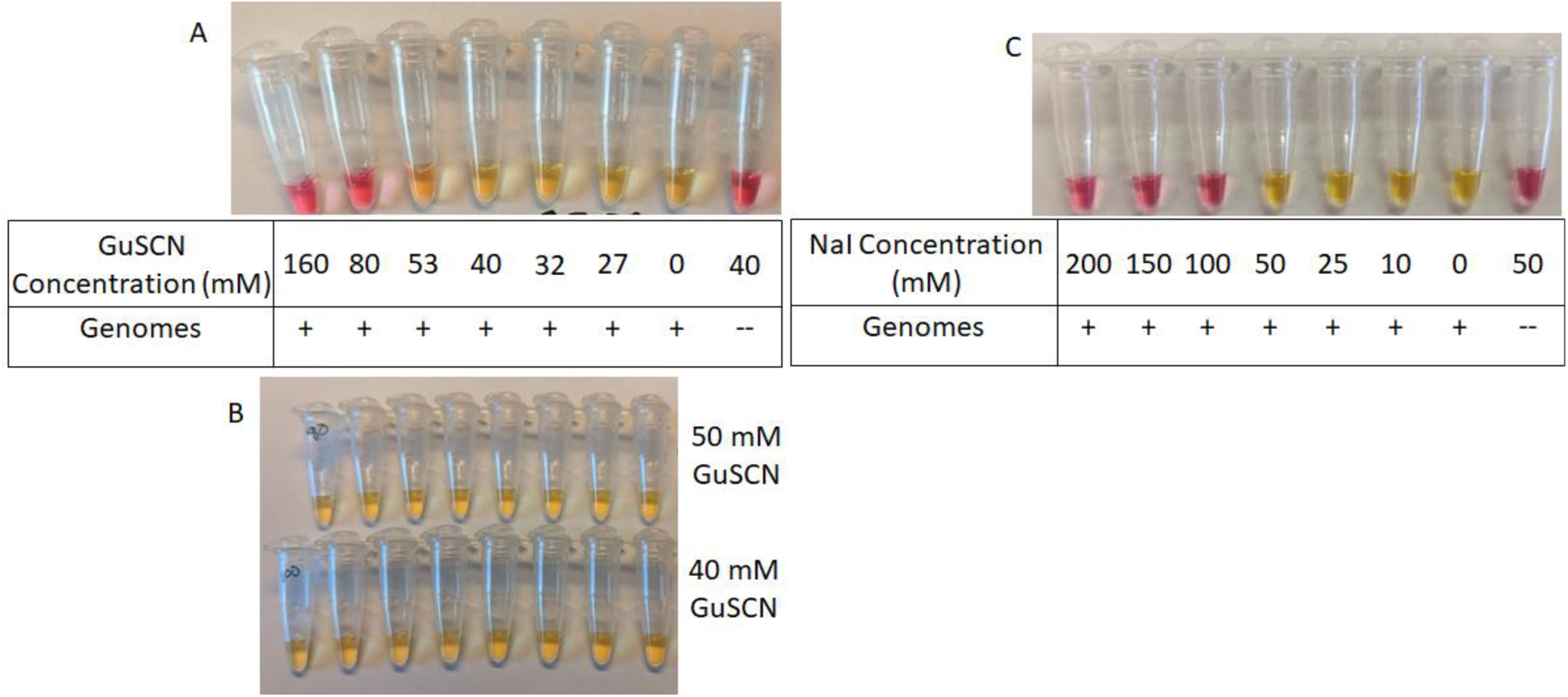
Assessment of RT-LAMP Chaotropic Salt Tolerance. 25 µl RT-LAMP reactions were run with a colorimetric readout. All reactions used the HMS Assay 1e primer set and 27 mM – 160 mM GuSCN (1:25 – 1:150 dilution of sample lysis buffer) or 10-200 mM NaI. Each reaction was incubated at 65°C for 30 minutes. A – Initial GuSCN range test, contained indicated GuSCN concentration and 500 control RNA copies (+) or 0 control RNA copies (--). B – Repeats of reactions with 500 control RNA copies and 50 mM of 40 mM GuSCN. C – NaI range test, contained indicated NaI concentration and 500 control RNA copies (+) or 0 control RNA copies (--).

Another chaotropic salt used in silica-based nucleic acid purifications is NaI^6^. We tested the tolerance of the RT-LAMP reactions with HMS Assay 1e primers in a similar fashion and found that at least 50 mM was tolerated (Figure 5C).

### Comparison of NEB and HMS Primer Sensitivity to Chaotropic Salts using Colorimetric Read-outs

While our final recommended protocol (below) does not utilize GuSCN, we did develop a protocol that does use it, for situations where GuSCN is the chaotropic agent in use. We tested different concentrations of GuSCN on the sensitivity of the RT-LAMP assay with various primer sets. This also served to directly compare the sensitivity of HMS Assay 1, HMS Assay 1e, NEB Gene N-A, and NEB Orf1a-C in the RT-LAMP reactions.

NEB Orf1a-C performed the worst following addition of GuSCN (Figure 6A), detecting 2/40 with 100 viral RNA copies and 5/40 with 200 viral RNA copies. This result was surprising, so we ran the experiment again, remaking all reagents including primer mixes and including four reactions of HMS Assay 1e with 200 viral RNA copies as a plate control (Figure 6B). The results were the same, with NEB Orf1a-C detecting 1/40 with 100 viral RNA copies, and 1-2/40 with 200 viral RNA copies (one of the 200 RNA copies reactions turned visibly orange, but not yellow). NEB Gene N-A performed better, detecting 11-13/40 with 100 viral RNA copies (depending on whether borderline orange reactions are considered to be positive) and 22-29/40 with 200 viral RNA copies (Figure 6C). HMS Assay 1 and HMS Assay 1e were much more sensitive. At 100 viral RNA copies, HMS Assay 1 and HMS Assay 1e detected 26/40 and 31/40, respectively (Figure 7). At 200 viral RNA copies, HMS Assay 1 and HMS Assay 1e detected 36/40 and 39/40, respectively. Furthermore, all positive reactions were completely yellow, leaving no ambiguous orange reactions. None of the reactions without viral RNA resulted in a positive reaction for any of the assays tested (Figures 6 and 7). There was also no difference in sensitivity between reactions that contained 50 mM GuSCN and those that didn’t when using the HMS 1e primers.

**Figure 6.**
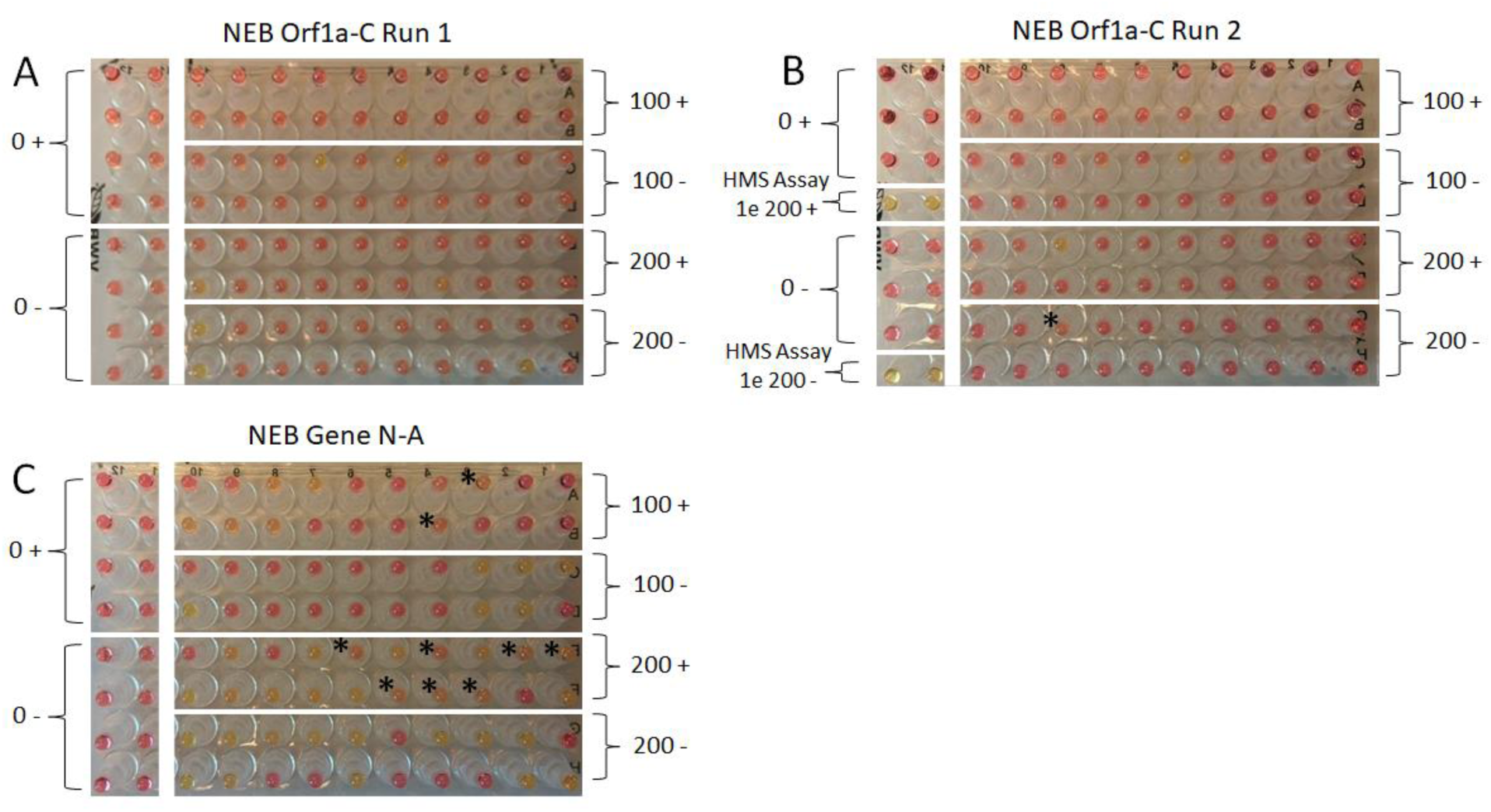
Assessment of GuSCN Effects on Sensitivity in NEB Colorimetric RT-LAMP Assays. 25 µl RT-LAMP reactions were run with a colorimetric readout. Number of control RNA copies per reaction (0, 100, or 200) noted. Reactions were run with 50 mM GuSCN (+) or without GuSCN (-) as noted. Each reaction was incubated at 65°C for 30 minutes. A – NEB Orf1a-C assay, first run. B – NEB Orf1a-C assay, second run. Note 4 HMS Assay 1e reactions run with 200 RNA copies with (+) and without (-) additional GuSCN as a plate control. C – NEB Gene N-A assay. * indicates reactions that were noticeably orange, but not completely yellow.

**Figure 7.**
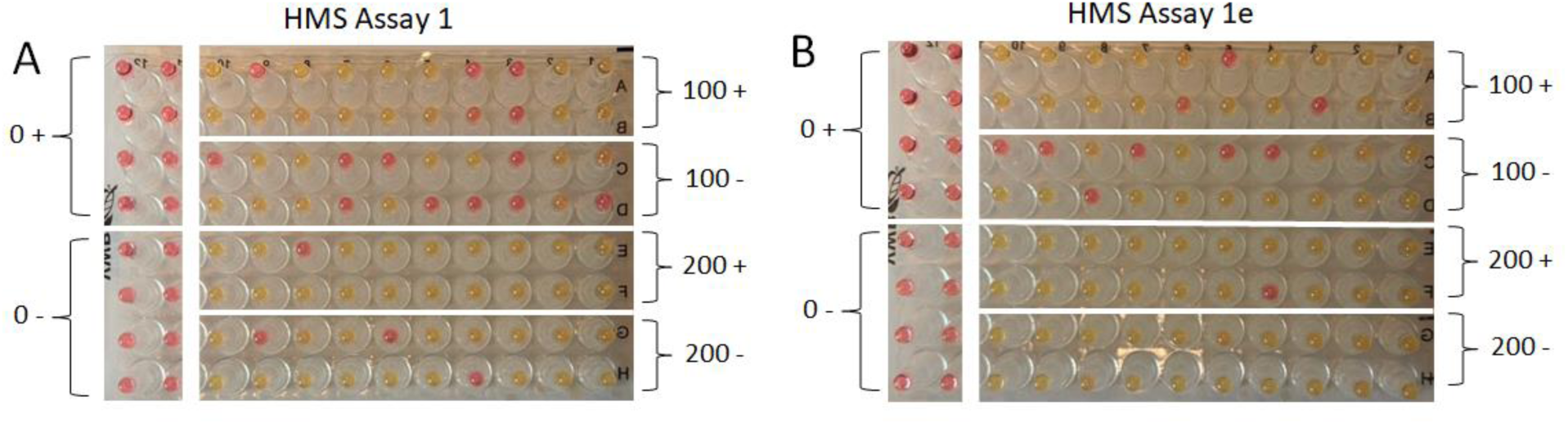
Assessment of GuSCN Effects on Sensitivity in HMS Colorimetric RT-LAMP Assays. 25 µl RT-LAMP reactions were run with a colorimetric readout. Number of control RNA copies per reaction (0, 100, or 200) noted. Reactions were run with 50 mM GuSCN (+) or without GuSCN (-) as noted. Each reaction was incubated at 65°C for 30 minutes. A – HMS Assay 1. B – HMS Assay 1e.

### Optimization of a Rapid Inactivation/Stabilization and Purification Protocol

The current sample collection methods used for SARS-CoV-2 testing require swabs to be placed in 2-3 ml of commercial collection media, such as Quest Diagnostics Viral Collection Media (VCM)^8^. This method presents a serious challenge for RT-LAMP-based detection as very little (no more than 1 µl) can be used in a 25 µl reaction due to the presence of dyes and buffers in the VCM that would prevent visualization of a pH shift in a positive reaction (data not shown). Other collection media, such as 0.9% saline, have less of an inhibitory effect, but the RNA in such media appears to degrade rapidly over the course of hours, reducing sensitivity if samples are not processed promptly. Finally, it is important to note that processing samples is not without risk, as such samples can contain infectious virus. Thus, we set out to design a protocol series that includes a rapid inactivation of virions in a variety of sample types while keeping the RNAse inactivated. In addition, our goal was to have this protocol enhance sensitivity, while being compatible with both direct addition to RT-LAMP reactions, and NaI-based purification (schematized in Figure 8).

**Figure 8.**
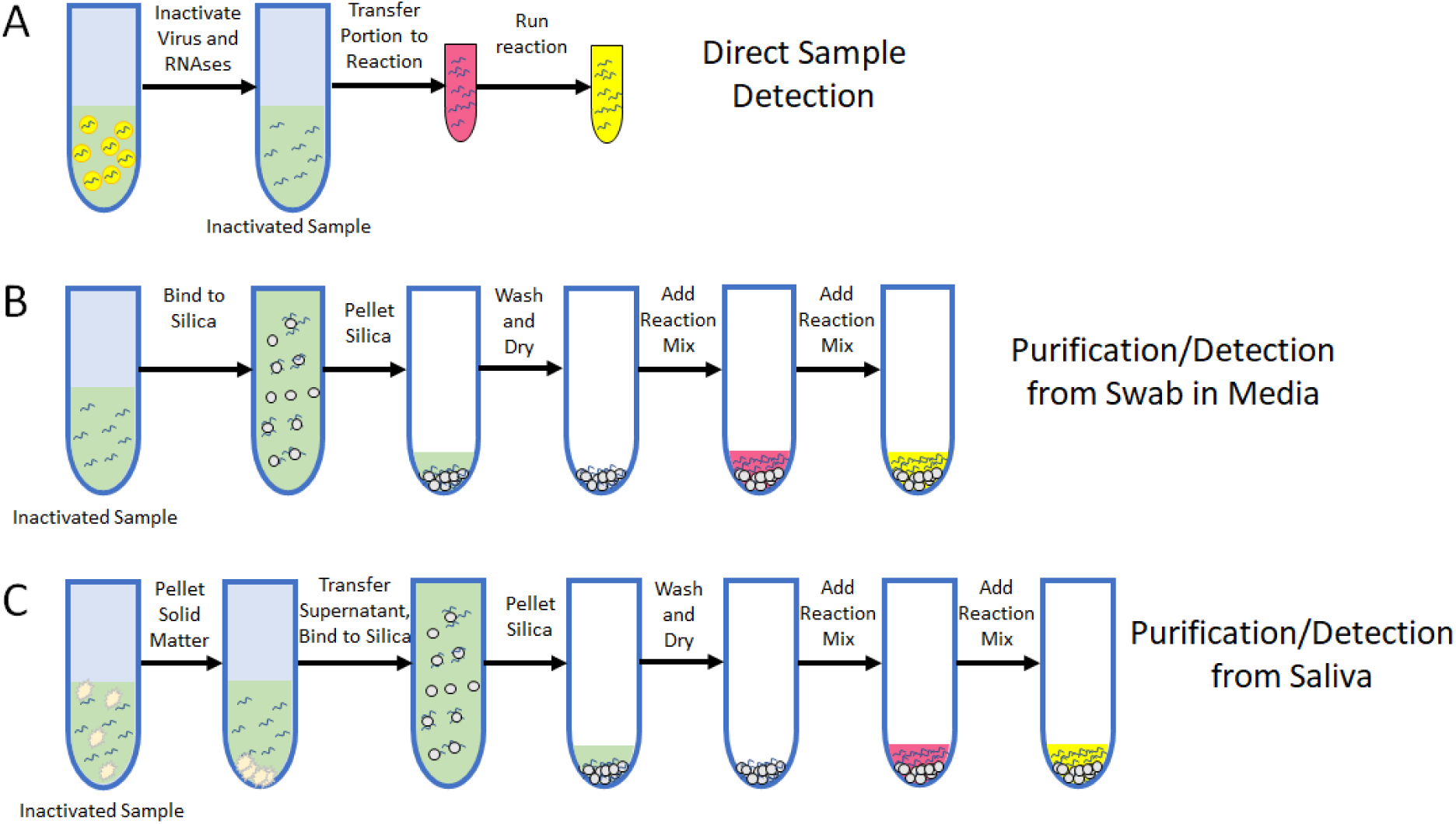
A Simple and Rapid Sample Inactivation and Purification Scheme. A – A schematic depicting sample inactivation and direct RT-LAMP testing. B – A schematic of the purification procedure for swab samples post inactivation depicting binding of viral RNAs to silica particles, washing away of impurities, and addition of a colorimetric RT-LAMP reaction. C – A schematic of the purification procedure for saliva samples post inactivation depicting clearing of the sample, binding of viral RNAs to silica particles, washing away of impurities, and addition of a colorimetric RT-LAMP reaction.

### Inactivation Protocol

In order to quickly inactivate/lyse virions while also protecting their RNA from endogenous RNAses, we employed a simple protocol utilizing a shelf-stable reducing agent, tris(2-carboxyethyl)phosphine (TCEP), the divalent cation chelator ethylenediaminetetraacetic acid (EDTA) and a brief period of heat (95°C). With this protocol, 1/100^th^ sample volume of a 100X TCEP/EDTA mixture (inactivation reagent) is added to the sample, which is then mixed and heated at 95°C for 5 minutes.

This protocol rapidly denatures proteins, utilizing the TCEP to reduce any disulfide bridges. Any divalent cations necessary for RNAse activity are released from denatured proteins and sequestered by the EDTA, rendering any renatured RNAses inert. We have found this sufficient to inactivate RNAse activity; an added benefit is that reducing agents have been shown to reduce the viscosity of saliva and mucus^9^. The resulting inactivated sample is fully compatible with direct addition to an RT-LAMP reaction, with at least 5 µl being tolerated. This protocol should be sufficient to inactivate any SARS-CoV-2 virions in the sample, rendering the sample much safer for downstream handling and transport. Previously developed lysis methods for RNA viruses use a 95°C lysis step, and MERS-CoV is highly sensitive to even 1 minute at 65°C^10, 11^.

Using this protocol, at least 5 µl of inactivated sample, including swabs in saline and 1XPBS, as well as straight saliva (without any other preparation), can be added to the reaction, allowing for robust detection at 50 RNA copies per microliter in the original sample (Figure 9). This protocol can also be completed with nothing more than this inactivation reagent and a boiling water source, allowing for rapid sample inactivation and stabilization in a wide variety of settings. The inactivation leaves a sample at a close to neutral pH with 2.5 mM TCEP and 1 mM EDTA, so such inactivated samples will likely be compatible with a variety of nucleic acid detection tests, not just RT-LAMP detection. We have also found dithiothreitol (DTT) can work in place of TCEP with a minor change in the sodium hydroxide (NaOH) used, as DTT is not as acidic as TCEP. DTT is far less stable, however, so we opted for TCEP as the optimal shelf-stable reagent.

**Figure 9.**
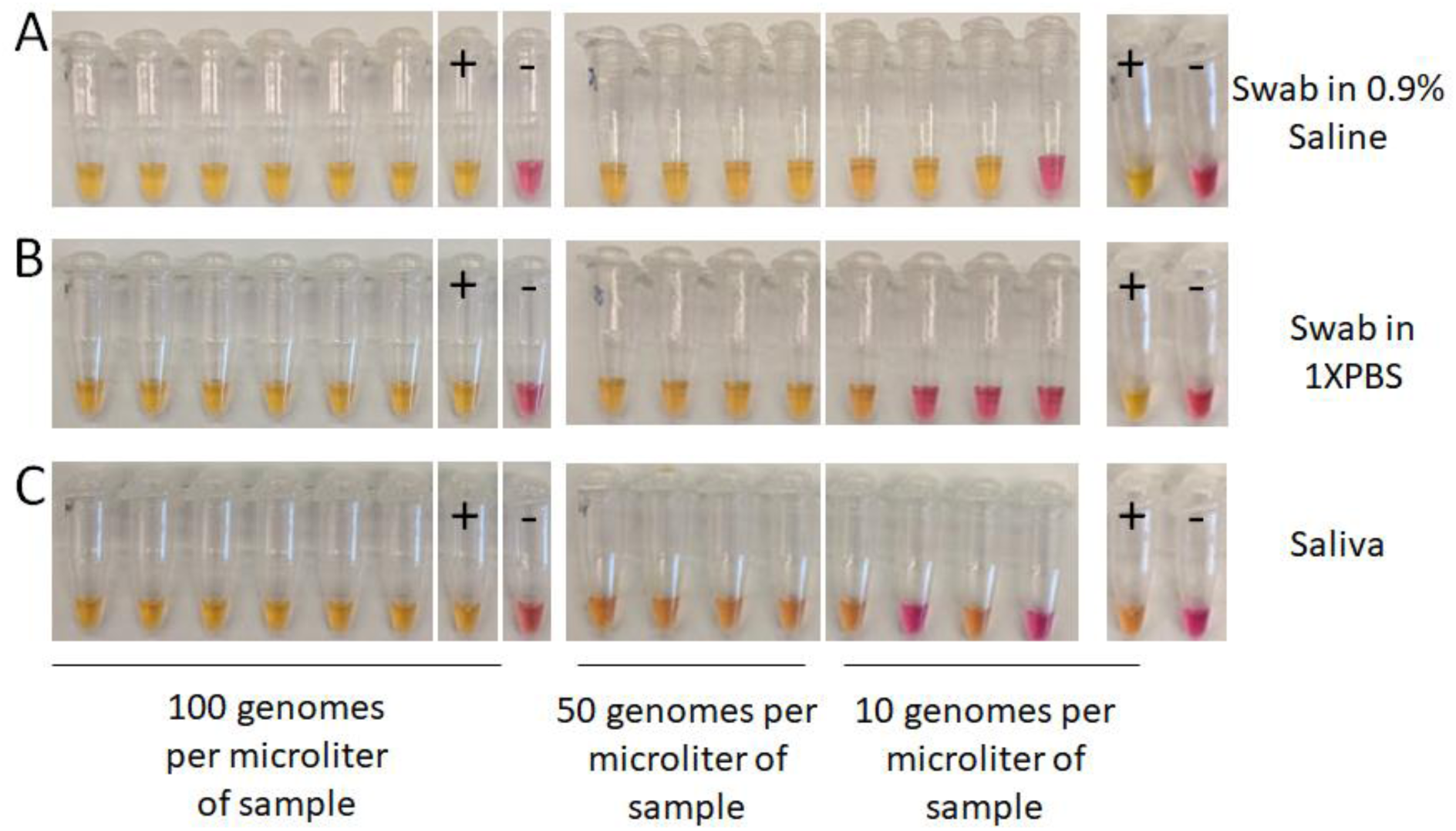
Sensitivity Test HMS Assay 1e Following Sample Inactivation. A-C - Sample inactivation was performed with mock sample throat and nasal swabs in saline (A) or 1XPBS (B) or straight saliva (C). Control viral RNAs were spiked into samples during inactivation once samples reached 95°C (to protect RNA from sample RNAse activity). Control RNA copies were added to the concentration indicated. Negative control samples (-) had no viral RNAs added. Following inactivation, 5 µl of sample was added to 20 µl of 1.25X HMS Assay 1e RT-LAMP reactions. Positive control reactions (+) had an additional 1000 control RNA copies added directly to the reaction. Reactions were run for 30 minutes at 65°C.

### Purification Protocol

In order to further increase the sensitivity of this test in a way that is inexpensive, accessible, and easily made to scale, we sought to optimize a purification protocol capable of concentrating viral RNA from a large sample into a single reaction. To do this, we used a very inexpensive and highly available silica particle suspension known as “glass milk”^6^. This suspension is made by cleaning small silicon dioxide particles and suspending them in an equal volume of water. A single lab can prepare enough for tens of millions of purifications in a day at a cost of less than $45 per 1 million purifications. This suspension was the forerunner to the commonly used silica-based column purification kits used today. As with these columns, nucleic acids will bind to the silica in the presence of chaotropic salts, such as GuSCN or NaI ^6, 7^.

Our preferred protocol for purifying RNA relies on NaI, which was often used as the chaotropic salt of choice for purifying DNA from agarose gels using glass milk^6^. In such protocols, three volumes of 6 M NaI was added to gel slices to melt the gel and bind the DNA to the glass milk. During our initial protocol optimizations using in vitro transcribed RNA to test different binding conditions, we found that RNA binds optimally when only a ½ sample volume of 6 M NaI is added (data not shown). This is quite convenient as using a smaller volume of a binding agent allows for a greater sample volume to be processed.

### Purification from Swabs in Saline/PBS

For samples that consist of swabs in saline or 1XPBS, one adds ½ volume of a NaI-based binding solution (6 M NaI, 2% TritonX100, 10 mM HCl) and 5 µl of glass milk to the sample following the previously described heat inactivation step (Figure 8B). The sample is then mixed and incubated at room temperature for 10 minutes to allow the RNA to bind (the samples are inverted every one to two minutes to keep the silica in suspension). The samples are then pulse spun in a tabletop microfuge, such as the VWR Galaxy mini (or in a larger tabletop centrifuge at 2,000 RCF), to pellet the silica, and the supernatant is poured off. A single wash with 80% ethanol is performed, with a final spin and use of a micropipette or fine tip transfer pipette to remove the last traces of 80% ethanol. The pellet of silica particles with bound RNA is dried, either at room temperature or at 65°C. Once dried, a 25 µl LAMP reaction can be added directly and the silica resuspended, at which point the RNA elutes into the reaction. The reaction can then be run directly, or the sample can be transferred to a different reaction vessel, with the silica particles included. As the reaction runs, the silica particles sink to the bottom of the tube and remain inert. When purifying from swabs in saline or 1XPBS, this allows for robust sensitivity down to at least 1 genome per microliter in the original sample (Figure 10A-B).

**Figure 10.**
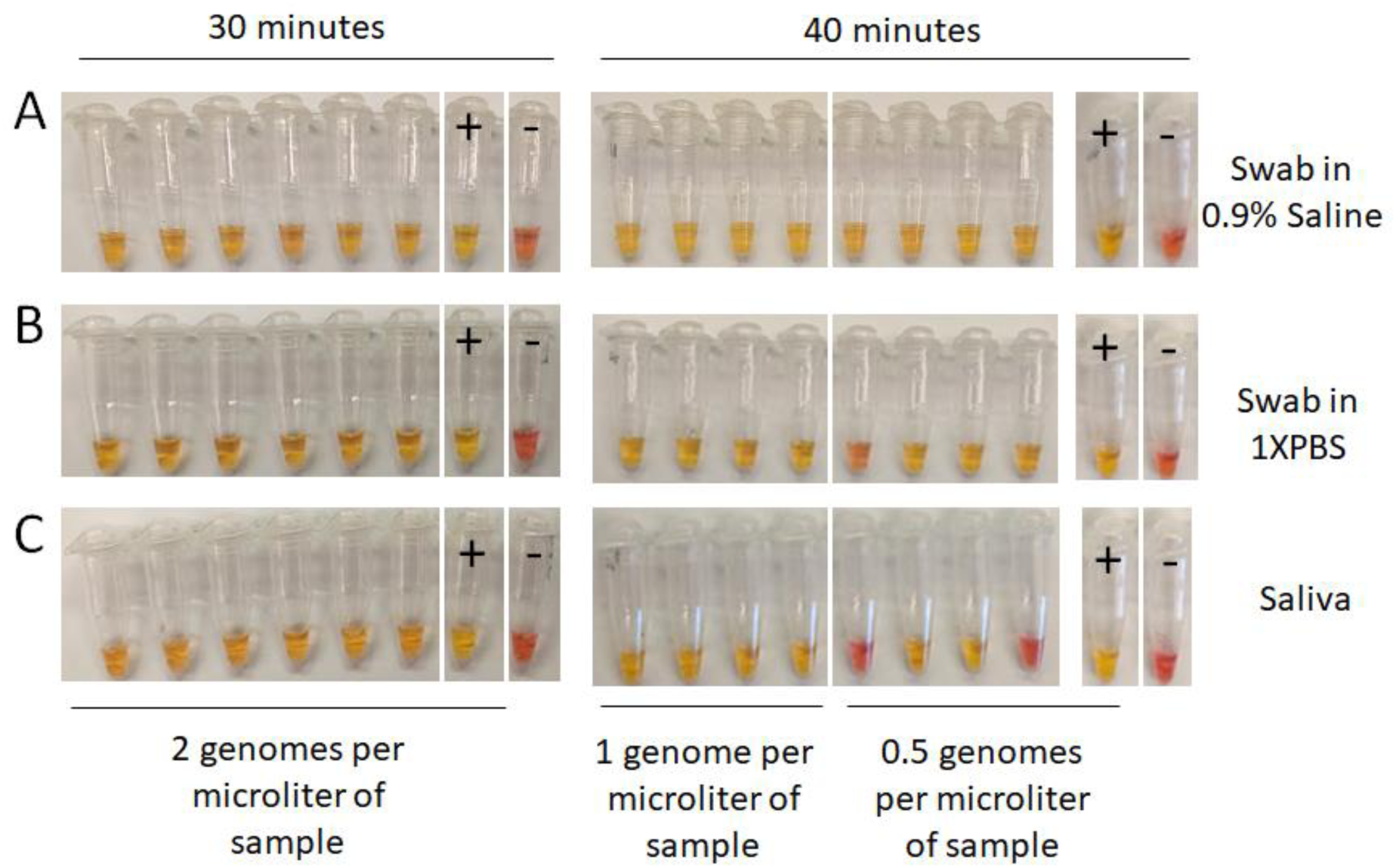
Sensitivity Test HMS Assay 1e Following Sample Purification. A-C - Sample inactivation was performed with mock sample throat and nasal swabs in saline (A) or 1XPBS (B) or straight saliva without swabs (C). Control RNA copies were spiked into samples during inactivation once samples reached 95°C. Control RNAs were added to the concentration indicated. Negative control samples (-) had no control RNA copies added. Following inactivation, samples were purified using glass milk with a centrifuge. 25 µl of 1X HMS Assay 1e RT-LAMP reaction was added to each dried pellet and the silica was resuspended. The entire slurry was then transferred to 0.2 ml PCR tubes. Positive control reactions (+) had an additional 1000 viral RNAs added directly to the reaction. Reactions were run for 30-40 minutes at 65°C, as indicated.

We have found that the protocol described above can be easily adapted to situations in which a centrifuge is not available. Following the binding of the RNA to the silica, samples can simply be allowed to sit undisturbed for 5-10 minutes, allowing the majority of the silica to settle to the bottom of the tube; the supernatant can then be poured off. For this procedure, we included two washes with 80% ethanol to ensure removal of inhibitors, allowing two minutes for the silica to sink to the bottom of the tube between each. We used a fine-tip transfer pipette to remove the final wash before drying the pellet and running the reaction as described above. While some silica particles are lost, particularly the smallest particles that don’t settle easily out of solution, we were still able to achieve a sensitivity down to at least 1 genome per microliter when purifying from 0.5 ml of swabs in saline of 1XPBS (Figure 11).

**Figure 11.**
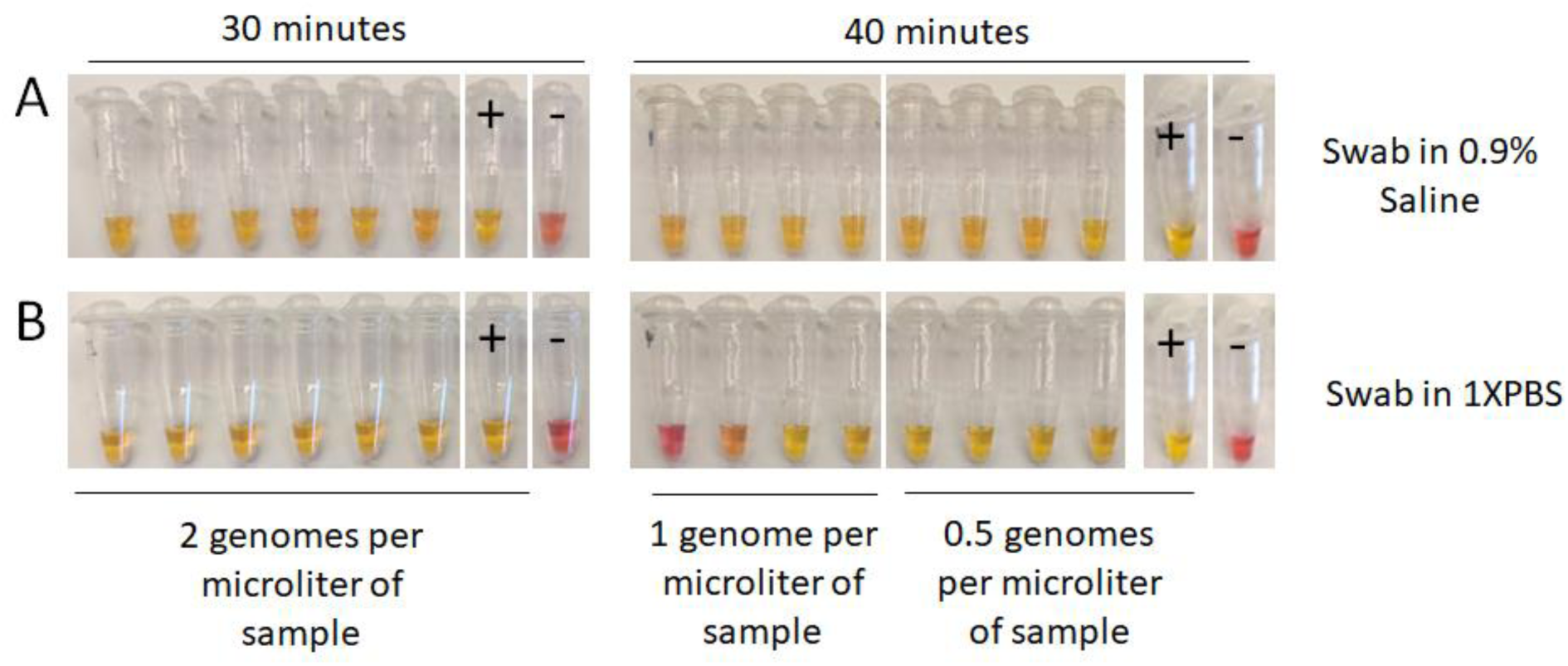
Sensitivity Test HMS Assay 1e Following Sample Purification without a Centrifuge. Sample inactivation was performed with mock sample throat and nasal swabs in saline (A) or 1XPBS (B). Control RNA copies were spiked into samples during inactivation once samples reached 95°C. Control RNA copies were added to the indicated concentration. Negative control samples (-) had no control RNA copies added. Following inactivation, samples were purified using glass milk without a centrifuge. 25 µl of 1X HMS Assay 1e RT-LAMP reaction was added to each dried pellet and the silica was resuspended. The entire slurry was then transferred to 0.2 ml PCR tubes. Reactions denoted by “+” had an additional 1000 control RNA copies added directly to the reaction. Reactions were run for 30-40 minutes at 65°C, as indicated.

### Purification from Saliva

We also sought to develop a protocol to purify viral RNA from saliva, as swab supplies are becoming limiting. We started with saliva inactivated by the inactivation protocol described above (Figure 8C). Before adding NaI or glass milk, the samples are spun in a benchtop microfuge, such as the VWR Galaxy mini, or another centrifuge, at 2,000 RCF for 10-15 seconds. This step causes the flocculant material in these samples to pellet, allowing the cleared sample to be poured into a fresh tube. Once this “pre-clearing” step is completed, the purification from the cleared supernatant can proceed exactly as described above for swab samples in media, allowing for a sensitivity down to at least 1 genome per microliter when starting with 0.5 ml of saliva (Figure 10C).

Unfortunately, we have been unable to perfect a protocol for saliva that does not rely on a small centrifuge. We tried allowing the flocculant material to pellet by gravity for 30 minutes (during which much of it does settle to the bottom of the tube, but not all) and transferring the rest to a fresh tube. This was unsuccessful, and even reactions which had positive control RNA copies spiked in directly before running failed to show a positive, indicating an inhibitor was still present (data not shown). We will continue to work on this, and invite others to do so as well, in order to make saliva-based purification protocols as accessible as possible.

### Purification from Commercial Collection Media

We also optimized protocols for purifying from commonly used collection media, including Quest Diagnostics VCM and PrimeStore MTM. Viral RNA can be purified from VCM following the heat-based inactivation with TCEP and EDTA, after which the gelatin in the collection media crashes out of solution. The sample can be spun down in a centrifuge and the cleared supernatant can be used for purification, with NaI binding solution supplemented with slightly more HCl. Samples in PrimeStore MTM can be treated in the same way as saline/PBS samples that have already been inactivated. Thus, ½ volume of the NaI binding reagent can be added directly and purification can proceed. With both, comparable sensitivities were seen compared with the other sample types described above (Figure 12).

**Figure 12.**
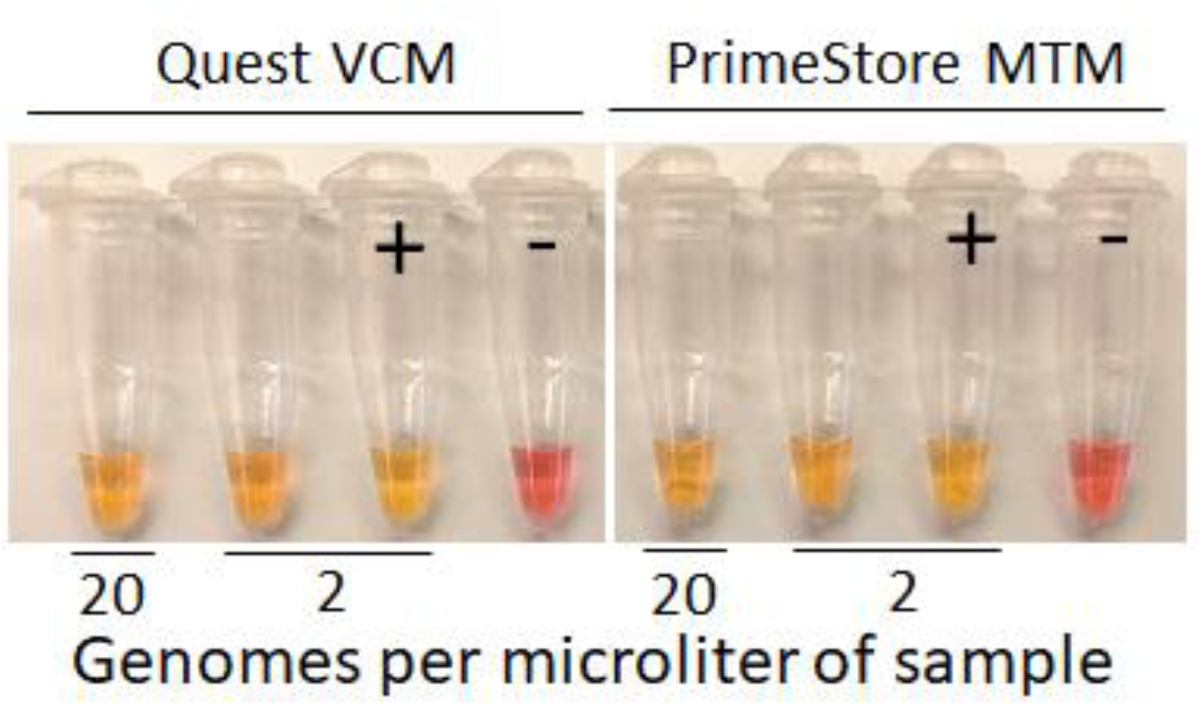
Purification from Commercial Collection Media. 750 µl of Quest VCM or 500 µl of PrimeStore MTM was spiked with viral RNA to the indicated concentration and purified as specified in Materials and Methods. Negative control sample (-) had no control RNA copies added to sample. Positive control reactions (+) had 1000 control RNA copies spiked directly into the reaction. Dried pellets were resuspended in 25 µl of 1X HMS Assay 1e RT-LAMP reactions and run for 30 minutes at 65°C.

### RNA Stability Post-Inactivation and Throughout Purification

In order to verify that endogenous RNAses are completely inactivated with these protocols, we inactivated samples of saliva, as well as swabs in saline or 1XPBS, and then added control viral RNA (100 copies per microliter) to probe for degradation. The inactivated samples with these control RNAs were incubated at 37°C for 30 minutes, giving any residual RNAse activity an opportunity to destroy control RNAs. When 5 µl of these samples were used in 25 µl RT-LAMP reactions with HMS Assay 1e primers, all reactions returned positive (Figure 13A). This indicates that RNAses are completely inactivated with this protocol. We also determined that the NaI concentration used during binding for purification (2 M) is sufficient to fully inhibit RNAse activity (Figure 13C-D). Finally, we found that, following purification, the dry RNA/silica pellet is quite stable. Following purification from 500 µl of swabs in saline inactivated with 2 RNA copies per microliter, the dried pellet was left at room temperature for 48 hours before adding 25 µl of RT-LAMP reaction with HMS Assay 1e primers. All reactions with viral RNA returned positive (Figure 13B).

**Figure 13.**
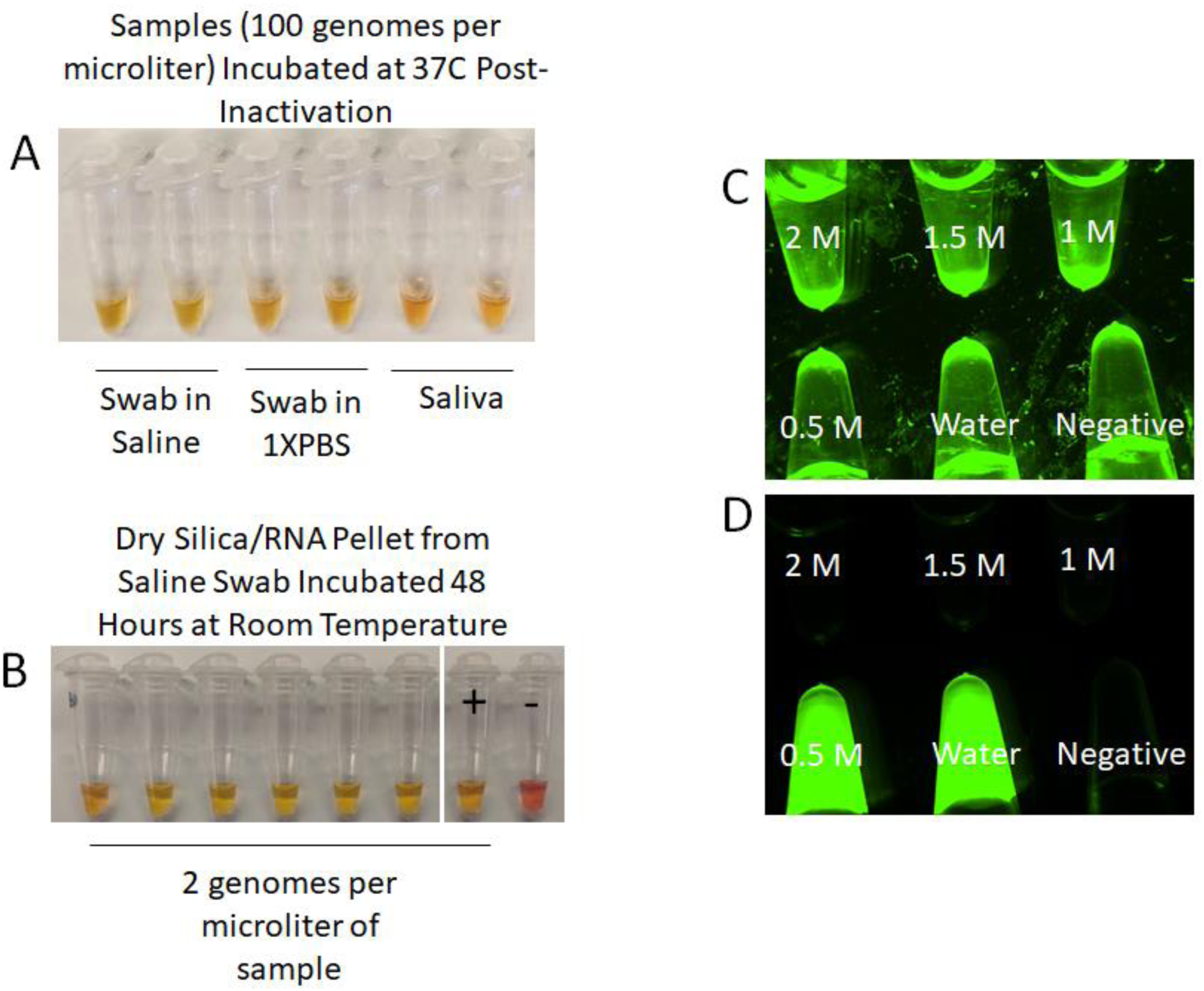
Evidence for RNA Stability from Inactivation Through Purification. A – Swabs in saline, swabs in 1XPBS, or saliva were inactivated and spiked with control RNA copies to 100 copies per microliter. After inactivation, samples were incubated at 37°C for 30 minutes before 5 µl of sample was added to 20 µl of 1.25X HMS Assay 1e RT-LAMP reactions which were run for 30 minutes at 65°C. B – 500 µl swabs in saline were purified using glass milk with a centrifuge. Once the silica/RNA pellet was dry, tubes were closed and left at room temperature for 48 hours before adding 25 µl of 1X HMS Assay 1e RT-LAMP reactions which were run for 30 minutes at 65°C. Negative control sample (-) had no control RNA copies added to sample. Positive control reactions (+) had 1000 control RNA copies spiked directly into the reaction. C-D – Swabs were placed in solutions of NaI, concentration indicated. 40 µl was used in a 50 µl RNAse Alert reaction and incubated for 30 minutes at 37°C. C – Brightfield. D – 488 nm fluorescence channel, fluorescence indicates RNAse activity.

## Discussion

In this report, we have presented evidence of a sensitive RT-LAMP assay for the SARS-CoV-2 virus made even more sensitive by a rapid and highly accessible inactivation and purification scheme. This scheme is compatible with current collection methods in which swabs are placed into a large volume of collection media, as well samples of straight saliva. This inactivation and purification scheme and RT-LAMP assay are simple and fast, relatively inexpensive, and do not rely on specialized equipment. Tests of these protocols in clinical settings are underway, with preliminary results looking promising (Anahtar et al., in preparation).

Our inactivation protocol allows for rapid inactivation of both virions and endogenous RNAses. This works both to stabilize viral RNA prior to detection and render samples safer for downstream handling. Furthermore, we see no reason that this inactivation protocol would be incompatible with most other nucleic acid detection assays, including the currently utilized FDA approved qRT-PCR test. Thus, this protocol may be suitable for a variety of testing pipelines to improve sample integrity and safety.

The silica particles used for purification are made from a crude silicon dioxide powder and can be prepared in enormous quantities very quickly, and very little is needed for each purification (one liter is enough for 200,000 purifications). A single lab can easily make enough for millions of tests, allowing for institutions with basic equipment like centrifuges and autoclaves to generate enough supply to meet high demand. Each swab sample purification can be performed in minutes in a single tube without a centrifuge, using only two solutions. Multiple purifications can be processed in parallel, allowing for efficient purification by medical personnel in point-of-care institutions.

These protocols can be used together in a single pipeline. Patient samples can first be inactivated and tested directly in an RT-LAMP assay with HMS Assay 1e primers, with a sensitivity of at least 50 viral RNA copies per microliter, in 30 minutes. Samples showing negative results in this assay can then be directly used in the glass milk purification protocol and retested, increasing the sensitivity to 1 viral RNA copy per microliter.

In practice, using multiple tests can ensure specificity and sensitivity. For example, if patient samples were tested separately with both HMS Assay 1e and NEB N-A assays, a double positive result would be a more certain positive result. Furthermore, if a genomic mutation were to prevent one assay from returning a positive result, a second assay targeting a different region of the genome could still detect the virus.

These inactivation and purification protocols are fully compatible with multiple tests. Inactivated samples can simply be added to multiple reactions. Following purification, the RNA/silica pellet can be resuspended in a larger volume of RT-LAMP mix without primers before dividing the resulting slurry among multiple tubes with different primer sets.

We understand that many rapid tests are being developed daily and are reaching FDA approval. However, given the incredible demand, a variety of tests with different components from different industry sources are needed to address the immediate shortage of tests in the face of a sweeping pandemic. Each protocol serves an important function, with different tests having different requirements, different sensitivities, and varying expenses.

## Materials and Methods

### Assay 1/1e Primer Design

The primary oligos for Assay 1/1e, F3, B3, FIP, and BIP primers, were designed by PrimerExplorer V5 (https://primerexplorer.jp/e/). The loop primers (LF and LB) were designed by hand, checking for appropriate melting temperatures using SnapGene software predictions.

### Oligos

All oligos were ordered from IDT and resuspended in UltraPure water at a 100 µM concentration. Oligos were combined to make 100 µl of 10X primer mix as follows – 16 µl FIP, 16 µl BIP, 2 µl F3, 2 µl B3, 4 µl LF, 4 µl LB, and brought to 100 µl with water.

### RT-LAMP Reactions

All RT-LAMP reactions were set up as described by NEB protocols (E1700 and M1800) and run at 65°C. Fluorescence based reactions were run as 10 µl reactions in a Bio-Rad CFX96 thermocycler for 60 minutes monitored every 30 seconds for fluorescence in the SYBR channel. Colorimetric assays were run as 25 µl reactions for 30 minutes at 65°C in an Eppendorf thermocycler, except for reactions following purification of samples with 0.5-1 viral RNA copies per microliter of sample, which were run for 40 minutes for improved color change. Colorimetric assays were imaged using a Pixel 2 smartphone with default settings.

### Control RNA

All viral RNA sequences used in this study were purified RNA controls from Twist Bioscience (SKU 102019, 1×10^6^ RNA copies per microliter), diluted appropriately in nuclease free water. They were non-overlapping RNAs representing fragments of the genome, as appropriate for each set of probes.

### Clean Reaction Setup

All reactions were assembled and sealed prior to running in a dedicated clean room that was regularly decontaminated with bleach and had limited personnel access. Once reactions were run, the reaction tubes or plates were never opened again to prevent post-amplification contamination of future reactions.

### Solutions

All solutions were created from molecular grade reagents. To make 5 ml of 100X inactivation reagent, first 358 mg of TCEP-HCl (Millipore Sigma 580567) was dissolved in water to create 2.5 ml of a 0.5 M solution. Then 1 ml of 0.5 M EDTA, pH = 8 (ThermoFisher Scientific AM9260G) was added. Finally, 10 N NaOH and UltraPure water (ThermoFisher Scientific 10977015) was added to bring the final volume to 5 ml and the NaOH concentration to 1.1-1.15 N NaOH (1.1 N for use with swabs in saline of 1XPBS, 1.15 N for use with saliva with a pH ∼6.5). For other collection media, the NaOH concentration will need to be optimized to ensure the pH of the final inactivated sample falls within an acceptable range such that the sample does not, upon addition, immediately cause the LAMP reaction to turn yellow or prevent the LAMP reaction from turning yellow upon successful amplification.

To make the NaI binding solution, 224.8 g of NaI (Millipore Sigma 793558) was dissolved in UltraPure water to a final volume of ∼230 ml. To this, 2.5 ml of 1 N HCl (made from 37% stock, Millipore Sigma 320331) and 5 ml of tritonX100 was added and mixed before bringing the volume to 250 ml with UltraPure water. Over time, this solution may turn somewhat yellow, presumably due to oxidation that results in the formation of molecular iodine. However, when added to an inactivated sample containing TCEP, this iodine is quickly reduced, rendering the solution colorless. This does not appear to affect the purification. Furthermore, this coloration can be inhibited by reducing the solution’s light exposure, e.g., wrapping the solution container in foil.

1XPBS was purchased as is (ThermoFisher 10010023) and 0.9% saline was created by bringing 1.54 ml of 5 M NaCl to 50 ml with UltraPure water.

### Glass Milk Preparation

To prepare glass milk, 325 mesh silicon dioxide (Spectrum Chemicals - SI108) was combined with an excess volume of 10% HCl (∼3 N HCl) made from combining 37% HCl (Millipore Sigma 320331) and MilliQ water (Millipore) in a fume hood (dry silica powder should not be inhaled). After acid washing for four to eight hours at room temperature, silica was pelleted by spinning two minutes at 5,000 xg and the supernatant was poured off. The pellet was resuspended in four pellet volumes of MilliQ water and then pelleted again. This wash step was repeated for a total of six washes. The pellet was then washed with four pellet volumes of 10 mM Tris HCl, pH = 8 (ThermoFisher Scientific AM9855G) and 1 mM EDTA (ThermoFisher Scientific 15575020), and pelleted. Finally, the pellet was resuspended in 1 pellet volume of 10 mM Tris HCl and 1 mM EDTA and autoclaved. This autoclave step is likely superfluous, however, as acid washes should render the beads free of contaminants. The resulting 50% glass milk slurry can be stored at room temperature. Before use, care must be taken to vigorously resuspend the particles as they begin to settle quickly.

### RNAse Activity Determination

RNAse activity was tested using IDT’s RNAseAlert substrate (IDT 11-04-02-03). Briefly, the detection substrate (an RNA oligo with a fluor and quencher) was resuspended in UltraPure water at a 10 µM concentration. For each test, 5 µl of this substrate and 5 µl of 10x buffer were combined with 40 µl of a solution that was being tested for RNAse activity. A test solution was created by submerging and vigorously agitating a cotton tip applicator (Puritan 806-WC) that had been swabbed thoroughly at the back of the throat, in 500 µl of the designated NaI solution. A positive control was created by submerging a swab in water, and a negative control had clean UltraPure water used without any additions. These reactions were then incubated for 30 minutes at 37°C and imaged in brightfield and 488 nm with a Leica stereoscope.

### Mock Samples

Mock swab samples were created in both saline and 1XPBS. To simulate a typical swab collection, one nasopharyngeal and one oropharyngeal swab were submerged and agitated in 3 ml of either solution. For saliva collection, first the mouth was rinsed with water. After 30 minutes without eating or drinking, saliva was collected.

### Sample Inactivation and Direct RT-LAMP Testing

All experiments with inactivated samples used either mock swab samples or saliva and RNA controls. To each sample, 1/100^th^ volume of 100X inactivation reagent was added. The samples were then mixed and placed in a heat block set to 95°C. After ∼2 minutes, by which point the samples reached 95°C, positive control RNAs were added and the remainder of the 5-minute inactivation proceeded. Samples were then cooled on ice. For testing remaining RNAse activity, samples, once cooled, were placed at 37°C for 30 minutes before being placed back on ice. For RT-LAMP testing, 5 µl of sample was added to 20 µl of 1.25X colorimetric RT-LAMP mix containing the HMS Assay 1e primer set.

### Sample Purification – with Centrifuge

All purification experiments used 500 µl of sample (either swabs or saliva) and were performed in 1.5 ml tubes (Fisher Scientific 14-222-155) whose conical shape made retention of the silica pellets very effective. Samples were inactivated as described above and cooled. For saliva purifications, these samples were spun at 2,000 xg in a VWR galaxy mini centrifuge for 10-15 seconds and the cleared supernatant was transferred to a fresh tube for purification. This step was omitted for swab samples. 250 µl of NaI binding reagent was added along with 5 µl of glass milk (these could be combined beforehand into a master mix format, 255 µl of which was added to each). The tubes were then mixed by inversion and incubated at room temperature for 10 minutes; they were inverted every two minutes to resuspend the silica. Samples were then spun for 2-3 seconds at 2,000 xg to pellet the silica, and the supernatant was poured off. 700 µl of 80% ethanol was added and the tubes were inverted 2-3 times to wash (the pellet need not be resuspended). Samples were spun again for 2-3 seconds and the supernatant poured off. Samples were spun for a final time to bring all residual 80% ethanol to the bottom and a micropipette or a fine tipped transfer pipette (such as Thomas Scientific 232-11) was used to remove the residual solution from the pellet. The pellet was then completely air-dried (until it resembled dry parchment) leaving the tubes open at room temperature or in a 65°C heat block for faster drying. 25 µl of 1X colorimetric RT-LAMP mix with HMS assay 1e primers was added and the pellet was resuspended by pipetting or flicking. The reaction can then be run directly by placing the tube at 65°C for 30 minutes (data not shown) or transferred to a 0.2 ml PCR tube before running at 65°C for 30 minutes (this tube format makes for easier imaging).

### Sample Purification – without Centrifuge

Purification using only gravity to pellet the silica particles was only successful using swabs, not saliva. 500 µl of sample was inactivated and RNA was bound to the silica with the NaI binding reagent as described above for use with a centrifuge. Samples were then allowed to sit undisturbed for 5-10 minutes to allow the silica to settle out, and the supernatant was poured off (some small particles remain in the supernatant which will be cloudy, but a significant amount of silica settled to the bottom of the tube). 700 µl of 80% ethanol was added and the tubes were inverted 2-3 times to wash (the pellet need not be resuspended). Samples were allowed to sit for 2-3 minutes and the supernatant poured off. An additional 700 µl of 80% ethanol was added and samples were allowed to sit for 2-3 minutes. The supernatant was then removed with a fine tipped transfer pipette moderately slowly (over 2-3 seconds) to leave as little 80% ethanol as possible. The pellet was then completely air-dried (until it resembled dry parchment) leaving the tubes open at room temperature or in a 65°C heat block for faster drying. Reactions were then run as described above for purifications with a centrifuge.

### Sample Purification with Commercial Collection Media

Viral RNA was spiked directly into clean samples of Quest Diagnostics VCM or PrimeStore MTM. For VCM samples (750 µl), 1/100^th^ volume of 100x inactivation reagent, described above, was added and samples were heated at 95°C for five minutes and cooled. VCM samples were then spun at 10,000 xg in a centrifuge for 15 seconds to pellet the gelatin at the bottom of the tube, and the cleared supernatant was transferred to a fresh tube (2/3 the original sample volume, 500 µl). ½ of this cleared supernatant volume of the NaI binding solution, with an additional 12.5 mM HCl, was then added, along with 5 µl of glass milk. Purification was then performed with a centrifuge as described above. For samples in PrimeStore MTM, no inactivation was performed. ½ sample volume of the NaI binding solution and 5 µl of glass milk was added and purification was then performed with a centrifuge as described above.

## Data Availability

All raw data (real-time thermocycler data or photographs) can be provided upon request.

## Acknowledgements

This work was supported by the Howard Hughes Medical Institute. We are grateful for the sharing of ideas and reagents with Nathan Tanner of NEB. Helpful discussions with Michael Springer of the Systems Biology Department of HMS are also gratefully acknowledged, particularly regarding the suggestion to use TCEP rather than DTT for the inactivation step. We were also guided in the development of these assays through helpful discussions with our colleagues, Melis Anahtar and Graham McGrath, of the Clinical Microbiology Department at the Massachusetts General Hospital. We would like to thank Jenne Etter, HMS Genetics Department Research Operation Manager, as well as HMS security, facilities, and delivery personnel for supporting our work during this challenging time. We would also like to thank Elizabeth Rabe and Joseph Rabe for their assistance in editing this manuscript.

## Notes

### Competing Interest Statement

Harvard University has filed patent applications regarding this technology on behalf of the inventors. Harvard believes strongly that while intellectual property rights serve to incentivize creation of new products, such rights should not become a barrier to addressing urgent and essential health-related needs. To address the global COVID-19 pandemic, Harvard is implementing technology transfer strategies to allow for rapid utilization of available technologies that may be useful for preventing, diagnosing and treating COVID-19 infection. More information can be found at otd.harvard.edu.

